# The impact of evaluation strategy on sepsis prediction model performance metrics in intensive care data

**DOI:** 10.1101/2025.02.20.25322509

**Authors:** Dang-Khoa Do, Patrick Rockenschaub, Sebastian Boie, Oliver Kumpf, Hans-Dieter Volk, Felix Balzer, Falk von Dincklage, Gregor Lichtner

## Abstract

**Background:** The prediction of the onset of sepsis, a life-threatening condition resulting from a dysregulated response to an infection, is one of the most common prediction tasks in intensive care-related machine learning research. To assess the performance of such models, different evaluation strategies (fixed horizon, peak score and continuous evaluation) are commonly employed, but there is no clear consensus on which approach should be used in order to provide clinically meaningful performance evaluation.

**Objective:** To assess different evaluation approaches of sepsis prediction models trained on a public intensive care dataset applied to German intensive care data.

**Methods:** In this retrospective, observational cohort study, we assessed the efficacy of machine learning models, pre-trained on the MIMIC-IV dataset, when applied to BerlinICU, a multi-site German intensive care dataset. To understand the real-world impact of implementing these models, we examined the performance variability across various evaluation strategies.

**Results:** The BerlinICU dataset includes 40,132 intensive care admissions spanning 10 years (2012-2021). Using the latest Sepsis-3 definition, we identified 4,134 septic admissions (prevalence 10.3%). Application of a temporal convolution network model to BerlinICU yielded an area under the receiver operating characteristic curve (AUROC) of 0.67 (95% CI: 0.66–0.68) for continuous evaluation with a 6-hour prediction horizon, compared to 0.84 (95% CI: 0.83–0.85) on the test set of MIMIC-IV. On BerlinICU, peak score evaluation showed a similar AUROC compared to continuous evaluation, while fixed horizon evaluation showed a reduced AUROC of 0.61 (95% CI: 0.60–0.62). Onset matching had minimal impact on performance estimates using continuous evaluation or fixed horizon evaluation but increased estimates for peak score evaluation. Performance metrics improved with shorter prediction horizons across all strategies.

**Conclusion:** Our results demonstrate that the choice of evaluation strategy has a significant impact on the performance metrics of intensive care prediction models. The same model applied to the same dataset yields markedly different performance metrics depending on the evaluation approach. Therefore, careful selection of the evaluation approach is essential to ensure that the interpretation of performance metrics aligns with clinical intentions and enables meaningful comparisons between studies. In our view, the continuous evaluation approach best reflects the continual monitoring of patients that is performed in real-world clinical practice. In contrast, fixed horizon and peak score evaluation approaches may produce skewed results when not properly matching the length of stay distributions between sepsis cases and control cases. Especially for peak score evaluation, longer visits tend to produce higher maximum scores because sampling from more values increases the likelihood of capturing higher values purely by chance.

## Introduction

Sepsis is a life-threatening condition resulting from a dysregulated response to an infection that leads to organ dysfunction (1), responsible for up to 20% of all deaths worldwide (2). Beyond its significant mortality rate (3), sepsis represents a tremendous financial burden, totaling to almost $24 billion dollars in the US in 2013 (4). Those who survive may experience long-term health consequences, including a diminished quality of life (5).

As each hour of delayed therapy increases mortality (6– 8), sepsis treatment guidelines continue to recommend early therapy within the first hour of sepsis recognition (9–11). Therefore, an early recognition of sepsis is essential. Numerous studies have investigated machine learning to aid early prediction of sepsis in the intensive care unit (ICU) (12, 13), including prospective application (14, 15) and external validation on data of different hospitals (15–17). Previous studies have employed different evaluation strategies, which vary in the criteria for data selection and lead to differences in the timing and scope of data considered relative to sepsis onset. These strategies can be broadly categorized into three groups:

### (I) Fixed Horizon Evaluation

This approach evaluates a model’s ability to predict the onset of sepsis at a predefined time point before the actual onset — the so-called prediction horizon — using all available data up to that time (13, 18). Varying the length of the prediction horizon allows for assessing the earliness of the model’s predictive ability. In this approach, it is particularly important to employ a case-control matching procedure, where control patients who do not develop sepsis are evaluated at a time point in their ICU stay that corresponds to the sepsis onset in sepsis patients. Without appropriate matching, control patients are typically assessed in the improved health conditions of their final pre-discharge hours. This comparison can oversimplify the analysis and bias results, as these conditions distinctly contrast with those of sepsis patients (19).

### (II) Peak Score Evaluation

Like fixed horizon evaluation, this approach evaluates a single prediction for each patient. However, instead of considering a pre-defined time interval before sepsis onset, this strategy utilizes the peak prediction score generated by the model across all time points during the patient’s stay before the actual onset (16). It has been reasoned that this approach better aligns with clinical practice, based on the assumption that a prediction model in a clinical setting operates with a fixed threshold and once any prediction score surpasses this threshold, an alarm for that particular patient is triggered (16, 20). Performance metrics derived under this evaluation strategy are considered to reflect the model performance in clinical practice. While excelling in simplicity, this strategy disregards temporal proximity of the prediction to the actual sepsis onset and favors extreme values, which might not be representative of the patient’s overall condition.

### (III) Continuous Evaluation

This approach evaluates a series of predictions made continuously for each patient over time (13). Specifically, predictions for all time points before sepsis onset are considered. In contrast to fixed horizon or peak score evaluation, which use a single prediction per patient with a label indicating whether the patient eventually develops sepsis, continuous evaluation assesses each time point individually, resulting in multiple predictions for every patient. All time points within the defined prediction window leading up to the onset are labeled “positive”, while time points before that window are labeled “negative”. The resulting performance metrics from this approach represent the model’s predictive efficiency across all time points, simulating its use for a randomly selected patient at a random time point. This view aligns well with clinical practice, where patients are continuously monitored without prior knowledge of when sepsis might occur (13).

In summary, fixed horizon evaluation assesses the model’s ability to predict sepsis onset at a specific, pre-defined time point before its actual occurrence. The peak score evaluation measures the model’s performance at the time point when the model outputs the highest probability for an upcoming sepsis event, reflecting a threshold-based clinical decision-making approach. The continuous evaluation approach evaluates the model’s predictive performance continually across all time points, reflecting the real-world scenario of continuous patient monitoring. It remains uncertain how the performance varies across these approaches and which approach most accurately reflects clinical practice.

We aimed to investigate how the choice of the evaluation approach influences the estimated performance of sepsis prediction models in an external validation context. Specifically, we compared the performance of models trained on a public intensive care dataset from the United States and subsequently applied it to German intensive care data. In doing so, we also address the lack of studies involving large-scale ICU cohorts from the German healthcare context, where the efficacy of sepsis prediction models—given the distinct patient demographics, treatment approaches, and healthcare policies— remains largely uninvestigated.

## Methods

### Datasets

We pre-trained models on the MIMIC-IV dataset, a well-known ICU dataset from the United States that is commonly employed to develop clinical prediction models (12, 13). We applied these models to BerlinICU, a comprehensive dataset from eight ICUs from Charité – Universitätsmedizin Berlin in Germany, one of Europe’s largest university hospitals. The ICUs were distributed across two different sites (Campus Virchow Klinikum and Campus Charité Mitte).

BerlinICU was extracted from the intensive care data management systems and the hospital information system present at the Charité – Universitätsmedizin Berlin. Specifically, we extracted the same clinical features that we used to pre-train the models, resulting in a total of 48 time-varying features, including vital parameters and laboratory results, along with 4 static features (age, sex, height and weight; Table S1). All clinical feature values outside of physiologically plausible ranges, as defined by the ricu package (21), were removed. The remaining data was aggregated into intervals of one hour using their median value. Missing values were imputed using a forward fill strategy. If no previous values were available, the training mean across all patients for that feature was used. Data were excluded from the dataset if they originated from non-intensive care wards, involved patients younger than 18 years, had a sepsis onset within 4 hours after admission, contained less than 6 hours of data or included gaps exceeding 12 hours. The dataset was restricted to data from the years 2012-2021 and limited to at most seven days following admission to the ICU (Figure 1), following the training setup (17).

**Fig. 1.**
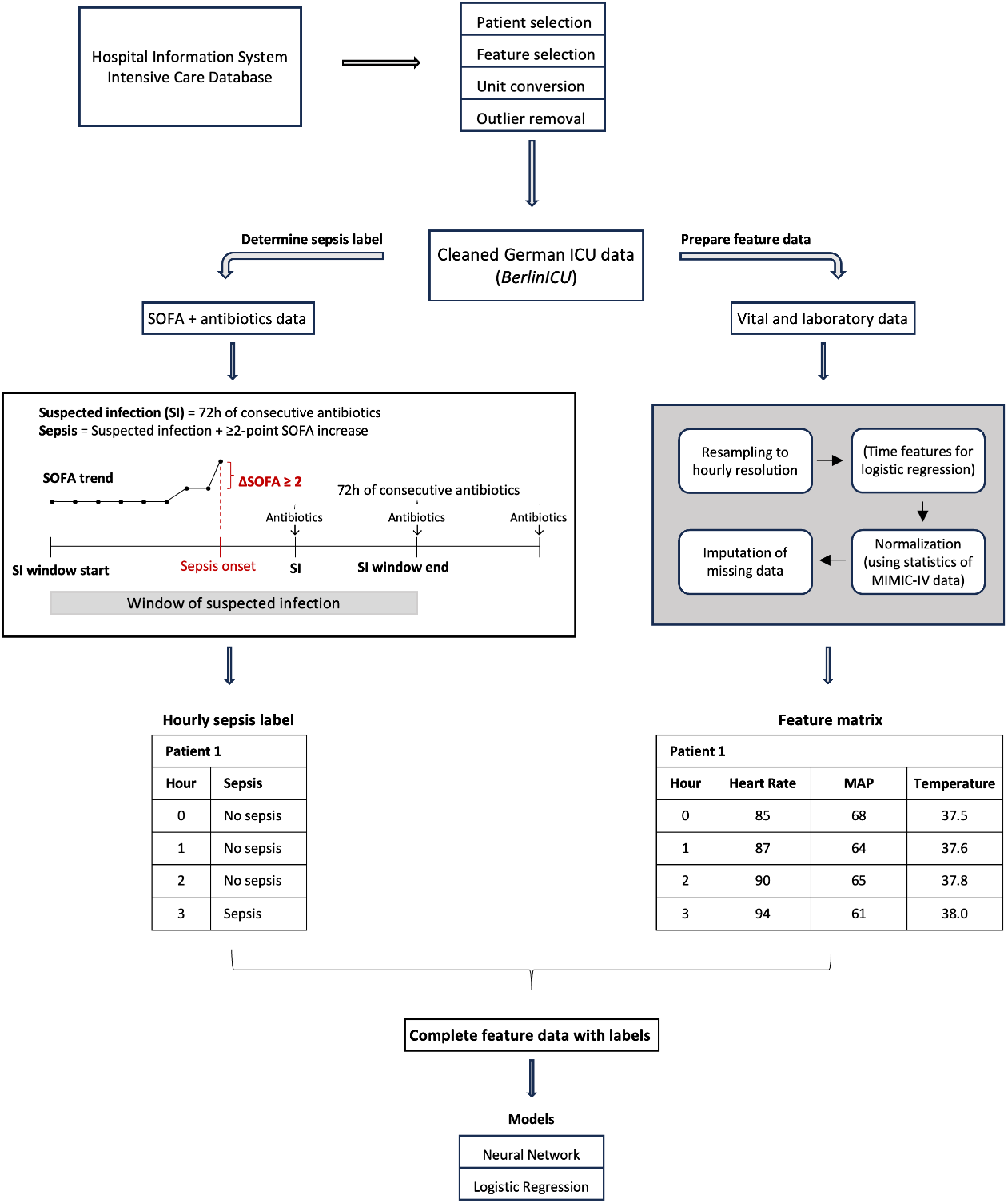
Preprocessing Pipeline. The BerlinICU dataset was extracted from the hospital information system and cleaned. We inferred sepsis onset according to the Sepsis-3-definition. For the logistic regression baseline models, we created additional temporal features. The final dataset was then passed to the models and evaluated. SOFA: Sequential Organ Failure Assessment score. MAP: Mean Arterial Pressure. ICU: Intensive Care Unit.

### Outcome definition

To identify the onset of sepsis, we used the latest Sepsis-3-definition criteria, defined as the cooccurrence of a suspicion of infection and an increase of the SOFA score by at least 2 points [1]. A suspicion of infection (SI) is present when antibiotic treatment is coupled with microbiological culture tests. As the BerlinICU dataset did not include microbiological data, we employed an alternative definition for SI similar to that followed in previous studies (16, 17): We inferred the SI from antibiotic treatment administered at least once every 24 hours for at least 3 days, with the SI start marked by the beginning of this treatment (Figure 1).

We assumed the onset of sepsis if an increase of at least 2 points of the SOFA score occurred within a 48-hour window preceding and a 24-hour window following the start of SI. The sepsis onset time point was defined as the time point of the recorded SOFA increase.

### Model training

We employed temporal convolutional network (TCN) models trained on the MIMIC-IV dataset, following the architecture and implementation described in (17). This choice is guided by a comprehensive study by Bai et al. (22), which demonstrated the superior performance of the TCNs in sequential data in various tests and domains. Our models were trained to predict, for each hourly bin, the individual risk of sepsis onset in the next 6 hours. We used a 10-times repeated random split scheme for hyperparameter tuning, resulting in 10 trained models for the hyperparameter set with the lowest average validation loss. For each repetition in this scheme, 20% of the data was held out as a test set. The remaining data was split into training (64%) and validation (16%). Training was performed using a binary cross-entropy loss and an Adam optimizer for a maximum of 1000 epochs with early stopping. To enhance model performance, binary missingness indicators were added for the 48 time-varying and 4 static features used as inputs.

For establishing baselines in our analyses, we utilized logistic regression models. These were trained following a 5-fold cross-validation scheme in which 80% of the data was used for training and 20% for testing. To better capture the temporal dynamics, we additionally engineered temporal features for the 48 time-varying features. These temporal features represent the trajectory of the measurements over time and include the minimum, maximum, mean, median and variance of the last 4, 8 and 16 hours for the 48 laboratory/vital features, totaling 772 features (48 time-varying features from the hourly bins, their 720 temporal features, 4 static features). To account for the class imbalance, we used class weights inversely proportional to class frequencies.

Training and evaluation were performed using python 3.10.8, pytorch 1.12.1 and scikit-learn 1.1.3.

### Model evaluation

To focus on evaluating the real-world applicability of the models in a clinical setting, we employed three different evaluation strategies that scrutinized different aspects of the models’ performance (Figure 2). For all strategies, we report the mean area under the receiver operating characteristic (AUROC). To estimate variability of performance metrics, we applied bootstrap resampling (1000 samples). For each evaluation setting that included a prediction horizon, we only included those patients whose length of stay matched or exceeded the defined horizon.

**Fig. 2.**
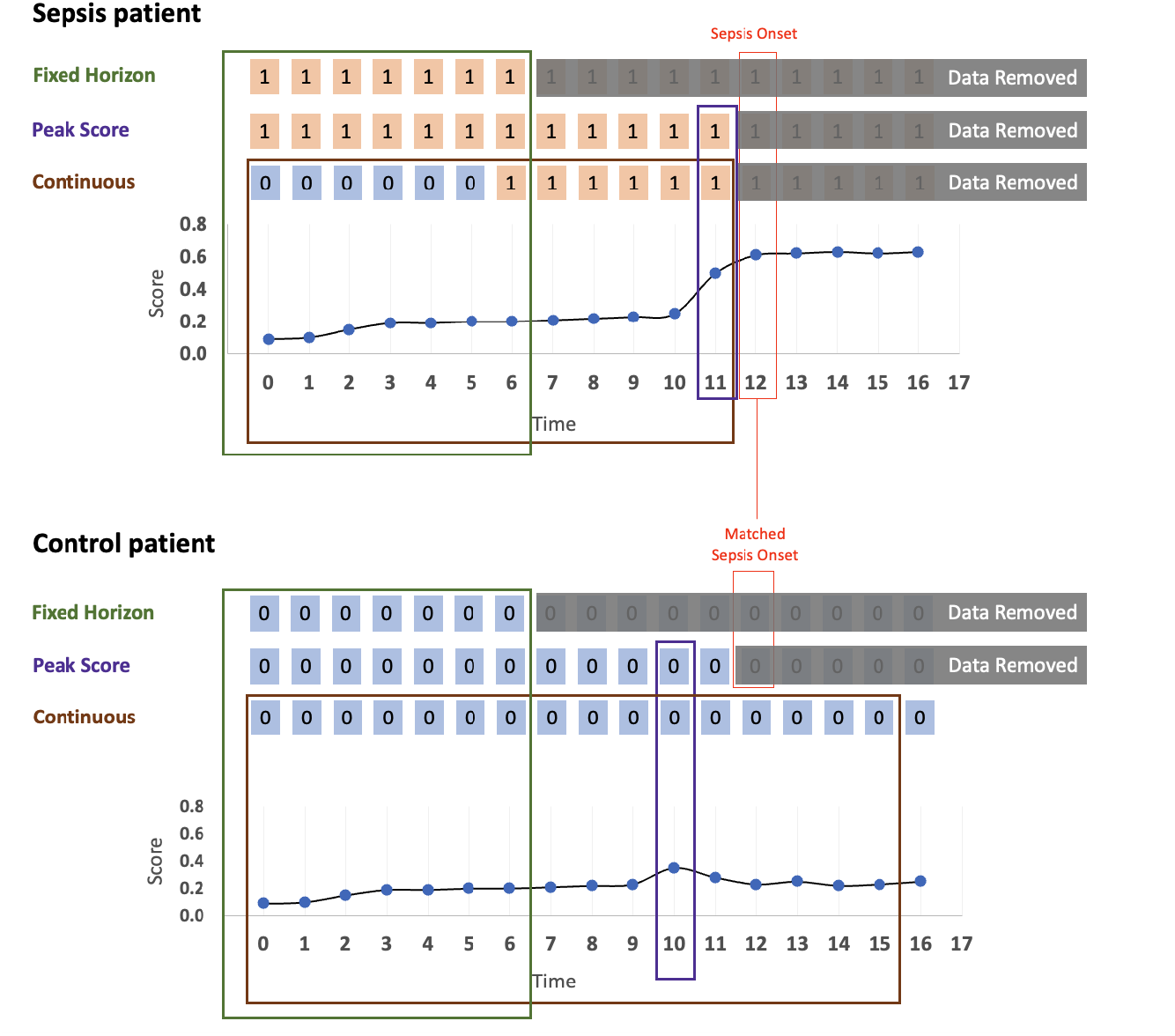
Overview of the evaluation strategies. For the **fixed horizon evaluation**, a prediction horizon was defined, where predictions were made for a time point that preceded the onset (or control onset) a pre-defined number of hours (6 hours in this example). Only the data from admission up to this time point was used to make the prediction (green rectangle). Labels were assigned as “positive” (marked as orange ‘1’s) for sepsis patients or “negative” (marked as blue ‘0’s) for control patients. In the **peak score evaluation**, the maximum prediction score across all time points prior to onset was determined (purple rectangle). Sepsis patients were labeled “positive”, while control patients were labeled “negative”. In the **continuous evaluation** (brown rectangle), for sepsis patients, time points within a specific time window before the onset (6 hours in this example) were labeled “positive”, while earlier time points – and all time points for control patients – were labeled “negative”. All time points before the onset (or control onset) were considered for evaluation. For **sepsis patients (top)**, the onset was the actual sepsis onset. For **control patients (bottom)**, the onset was defined either as a hypothetical control onset matched to the onset time of a sepsis patient (“matched onset”) or as the last available data point, either at discharge or at 7 days after admission, whichever occurred first. The rectangular frames represent the data that was ultimately included for each evaluation strategy. Shown here are the default settings for each evaluation strategy: the fixed horizon and peak score evaluations use matched sepsis onset times, while the continuous evaluation is depicted without onset matching.

A key aspect of the evaluation strategies is the handling of time series data for non-septic patients (controls): While septic patients (cases) have a clearly defined sepsis onset that marks the endpoint of their time series for analysis, non-septic patients lack such a reference point. Therefore, we employed two commonly used approaches to define the end-point of their time series for analysis:

#### No Onset Matching

The hypothetical onset for control patients is defined as the last available data point, either at discharge or at 7 days after admission, whichever occurred first.

#### Onset Matching

Onsets of control patients are aligned with those of septic patients by randomly pairing each control patient with a sepsis patient whose time to sepsis onset does not exceed that of the control patient. The onset time of the matched sepsis patient is then used as the hypothetical onset time for the control patient (16, 19, 23).

### Fixed horizon evaluation

Fixed horizon evaluation uses all data up to a certain time point before sepsis onset, known as the prediction horizon (*h*), to predict the upcoming onset of sepsis. Here, the predicted score *y*_*pred*_, which is based on all available data for the patient up to *h* hours before the sepsis onset for sepsis patients or up to a hypothetical onset for control patients (*t*_*onset*_), and the true label *y*_*true*_ are given by:

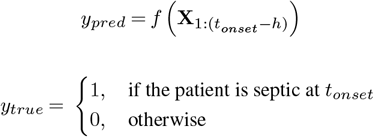

Where **X**_1:*t*_ is the patient data (feature matrix) from time point 1 to *t* and *f* is the model that outputs a score correlated to the probability of a sepsis onset.

### Peak score evaluation

To evaluate the model’s performance throughout the entire patient encounter, the maximum model prediction score from ICU admission to the onset of sepsis or control offset was determined, resulting in a single score/label pair for each patient for performance score calculation. This method focuses on the peak confidence of the model for sepsis prediction during a patient’s stay. Here, the prediction score and true label are given by:

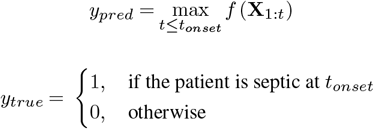

For horizon-dependent analysis, we used the peak score within the defined horizon instead of across all time points of each patient.

### Continuous evaluation

To evaluate the model’s capability to predict the onset of sepsis at a randomly selected time point for a randomly selected patient, we implemented a continuous horizon-based prediction strategy. This included first establishing a specific time horizon (e.g., 6 hours) to assess the model’s ability to predict the sepsis onset at any given time point within that timeframe. For sepsis patients, a positive label was assigned to all time points within the horizon, while all other time points were labeled as negative. For control patients, all time points were labeled as negative. We included the prediction scores from each hourly interval up to the onset of sepsis in sepsis patients or the hypothetical control onset in control patients (*t*_*onset*_), excluding the prediction at the actual onset (or control onset). The prediction score and true label are given by:

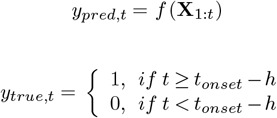

This procedure generated a sequence of hourly score/label pairs for each patient:

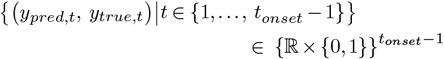

To accommodate variations in the length of these sequences among patients, we applied inverse frequency weighting to the samples when computing performance metrics.

### Ethics approval

This study was approved by the local ethics committee (Ethikausschuss am Campus Virchow-Klinikum, Charité—Universitätsmedizin Berlin, Chairperson PD Dr. E. Kaschina, Application Number EA2/137/22, approval date: 12 Jul 2022, amendment date: 10 May 2023).

## Results

### Study cohort

From 60,332 ICU stays in the initial dataset, after extraction, preprocessing and filtering the final dataset consisted of 40,132 ICU stays of 36,872 unique patients spanning 10 years (2012-2021). 4,134 stays were septic, resulting in a sepsis prevalence of 10.3% (Figure 3, Table 1). The original MIMIC-IV training dataset consisted of 67,056 ICU stays with a sepsis prevalence of 5.6% (3,730 ICU stays).

**Table 1.** Patient characteristics of BerlinICU and MIMIC-IV for the total dataset, the sepsis cohort and non-sepsis cohort.

| Statistic | Total | Sepsis | Control |
| --- | --- | --- | --- |
| **BerlinICU** |  |  |  |
| Cohort size (n (%)) | 40,132 (100%) | 4,134 (10%) | 35,998 (90%) |
| Age (years; Median (IQR)) | 64 (50-75) | 67 (55-77) | 63 (50-75) |
| ICU length of stay (days; Median (IQR)) | 1.0 (0.7-2.1) | 2.0 (1.4-3.4) | 1.0 (0.7-2.0) |
| Sex, male (n (%)) |  |  |  |
| Male | 22,423 (56%) | 2,444 (59%) | 19,979 (56%) |
| Female | 17,709 (44%) | 1,690 (41%) | 16,019 (44%) |
| Patient counts (%) |  |  |  |
| Patients on vasopressors | 17,117 (43%) | 3,442 (83%) | 13,675 (38%) |
| Patients ventilated | 13,967 (35%) | 2,774 (67%) | 11,193 (31%) |
| Patients on any antibiotics | 22,155 (55%) | 4,134 (100%) | 18,021 (50%) |
| Patients on antibiotics for ≥ 3 days | 5,351 (13%) | 4,134 (100%) | 1,217 (3%) |
| **MIMIC-IV** |  |  |  |
| Cohort size (n (%)) | 67,056 (100%) | 3,730 (5.6%) | 63,326 (94.4%) |
| Age (years; Median (IQR)) | 65 (53-76) | 67 (54-76) | 65 (53-76) |
| ICU length of stay (days; Median (IQR)) | 1.7 (1.0-2.8) | 0.8 (0.6-1.5) | 1.7 (1.0-2.8) |
| Sex, male (n (%)) |  |  |  |
| Male | 37,184 (55%) | 2,132 (57%) | 35,052 (55%) |
| Female | 29,872 (45%) | 1,598 (43%) | 28,274 (45%) |
| Patient counts (%) |  |  |  |
| Patients on vasopressors | 8,548 (13%) | 1,230 (33%) | 7,318 (12%) |
| Patients ventilated | 21,863 (33%) | 2,520 (68%) | 19,343 (31%) |
| Patients on any antibiotics | 46,276 (69%) | 3,730 (100%) | 42,546 (67%) |
| Patients on antibiotics for ≥ 3 days | 6,214 (9%) | 2,760 (74%) | 3,454 (5%) |

**Fig. 3.**
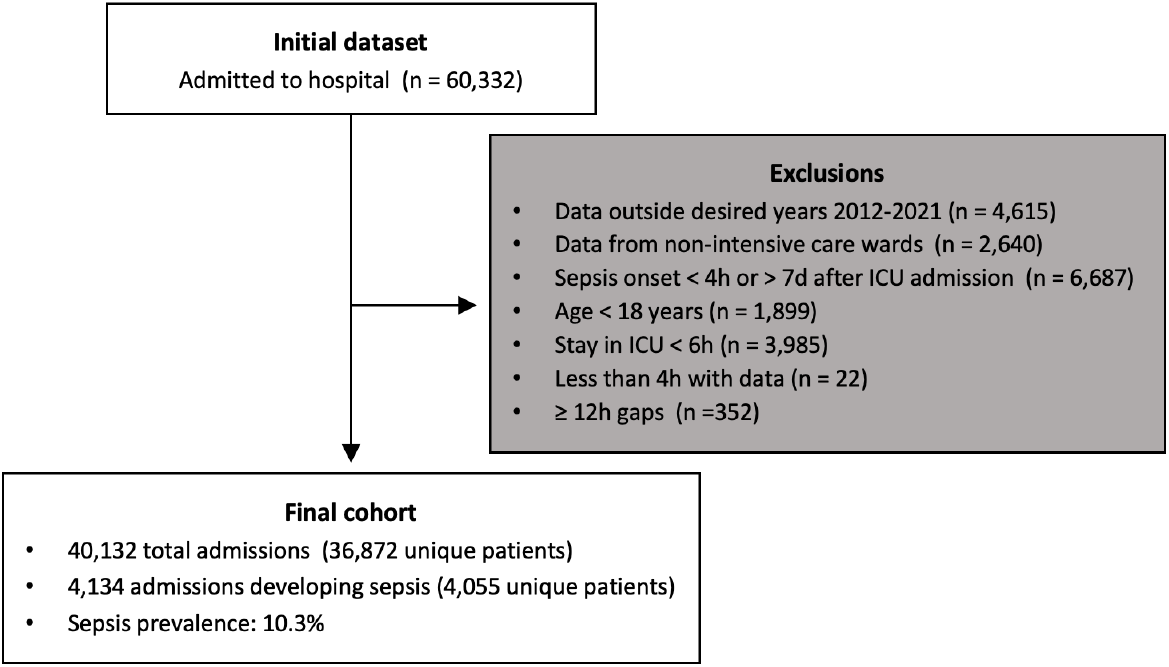
Flow diagram of patient eligibility criteria.

### Influence of evaluation strategy on model performance estimates

Applying the TCN models pre-trained on MIMIC-IV to the MIMIC-IV test set for internal validation under a scenario aligned with the training setup - continuous evaluation using a 6-hour prediction horizon without onset matching - resulted in an AUROC of 0.84 (95% CI: 0.83-0.85, Figure 4a). When the same models were applied to BerlinICU for external validation under the same evaluation conditions, performance dropped to 0.67 (95% CI: 0.66-0.68, Figure 4b).

**Fig. 4.**
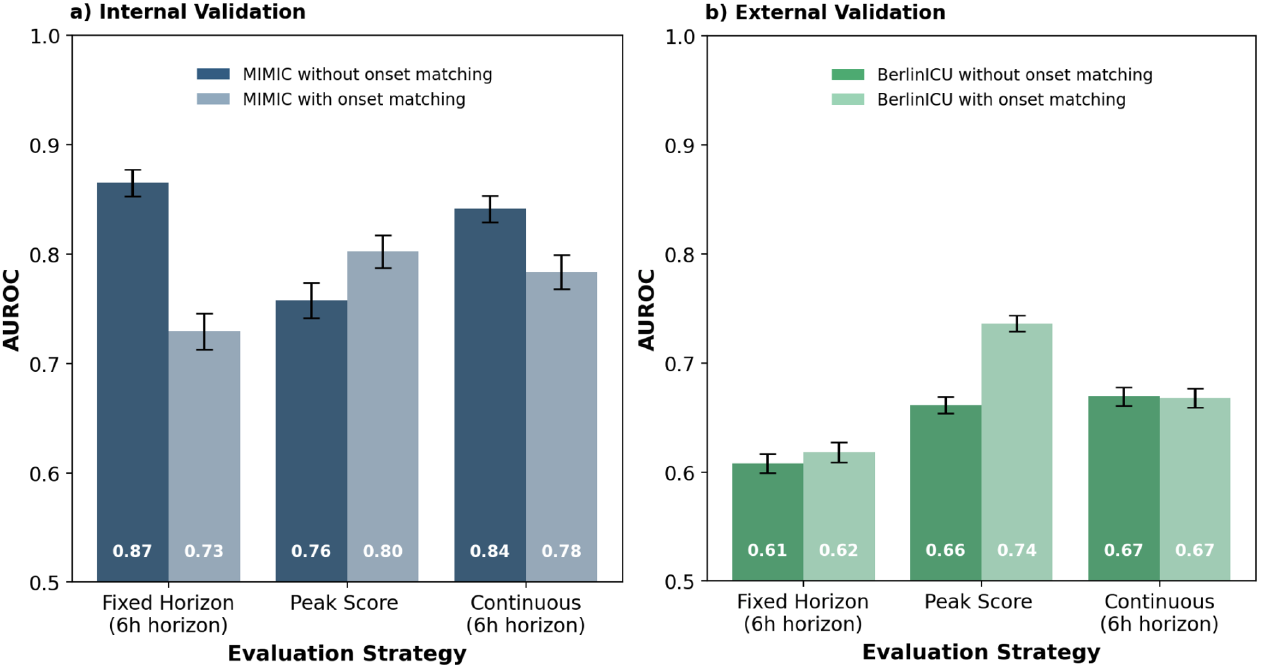
Comparison of model performance of temporal convolutional network (TCN) models for different evaluation strategies. The models were applied to the test splits of the MIMIC-IV training dataset (a) and the entire German ICU dataset (b; BerlinICU). Model performance was evaluated using fixed horizon evaluation (6h horizon), peak score evaluation (entire patient stay) and continuous evaluation (6h horizon) without onset matching (dark bars) and with onset matching (light bars). The AUROC value inside the bar represents the mean AUROC across all bootstrap samples and models. AUROC: area under the receiver operating characteristic. Error bars: 95% confidence intervals.

To assess the impact of different evaluation strategies, we performed fixed horizon evaluation under the same conditions (6-hour prediction horizon, no onset matching) and peak score evaluation, which does not employ a prediction horizon. On the MIMIC-IV test set, fixed horizon evaluation resulted in a slightly higher AUROC of 0.87 (95% CI: 0.85-0.88), whereas peak score evaluation showed significantly reduced performance of 0.76 (95% CI: 0.74-0.77). In external validation on BerlinICU, fixed horizon evaluation resulted in a significantly reduced AUROC (0.61 [95% CI: 0.60-0.62]) compared to continuous evaluation (ΔAUROC −0.06), while the peak score evaluation estimate (0.66 [95% CI: 0.65-0.67]) was similar to that of the continuous evaluation (ΔAUROC −0.01). Across all evaluation strategies, performance estimates were consistently lower on BerlinICU compared to the MIMIC-IV test set.

Aligning the length of stay distribution of control patients to sepsis patients through onset matching led to a notable AUROC reduction in internal validation on the MIMIC-IV test set for continuous evaluation (ΔAUROC −0.06) and fixed horizon evaluation (ΔAUROC −0.14), while AUROC estimates for peak score evaluation increased (ΔAUROC 0.04). On BerlinICU, onset matching had minimal impact on performance estimates during continuous evaluation or fixed horizon evaluation. In contrast, for peak score evaluation, onset matching increased AUROC estimates (ΔAUROC 0.08).

Across all evaluation strategies, TCN models consistently outperformed logistic regression baseline models (Figure S1). The only exception was observed in fixed horizon evaluation on BerlinICU without onset matching, where logistic regression showed a superior performance (AUROC 0.64 [95% CI: 0.63–0.65]) compared to TCN (AUROC 0.61 [95% CI: 0.60–0.62]).

### Influence of the prediction horizon on model performance estimates

To evaluate the ability of the models to provide timely predictions, we assessed their performance across a range of prediction horizons (Figure 5). Performance estimates generally improved with shorter prediction horizons across all evaluation strategies. During internal validation on the MIMIC-IV test set, fixed horizon evaluation and continuous evaluation showed consistent improvement, with the highest AUROC achieved at a 1-hour horizon (fixed horizon: AUROC 0.92 [95% CI: 0.91-0.93], continuous: AU-ROC 0.87 [95% CI: 0.85-0.88]). Peak score plateaued around the 6-hour horizon (AUROC 0.92 [95% CI: 0.91-0.93]), with only marginal improvements for smaller horizons. In external validation on BerlinICU, performance also generally increased with shorter prediction windows, with few exceptions including a plateau around the 6-hour horizon in fixed horizon and peak score evaluation with onset matching.

**Fig. 5.**
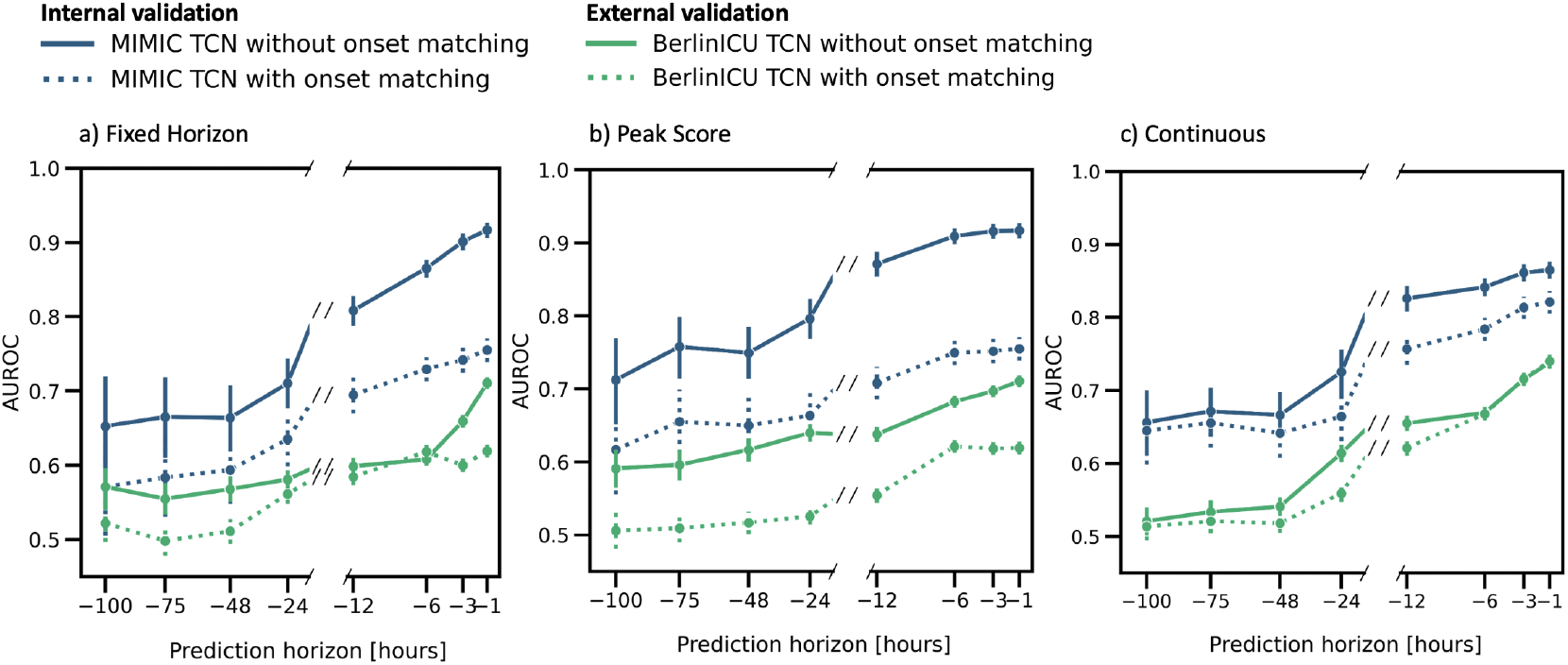
Performance metrics across prediction horizons, comparing the performance of temporal convolutional network (TCN) models for an evaluation with onset matching (dashed lines) and without onset matching (solid lines). The models were applied to the test splits of the MIMIC-IV training dataset (blue) and the entire German ICU dataset (BerlinICU; green). Shown is the AUROC based on varying prediction horizons from one hour before onset up to 100 hours before onset. AUROC: area under the receiver operating characteristic. Error bars: 95% confidence intervals.

Onset matching consistently decreased performance estimates during internal validation on the MIMIC-IV test set across all evaluation strategies, albeit to different extents, with continuous evaluation showing the smallest susceptibility to the matching approach. On BerlinICU, onset matching impacted strategies differently depending on the prediction horizon. Notably, continuous evaluation remained more robust to onset matching, maintaining more consistent AU-ROCs compared to the other strategies across different prediction horizons, particularly for very short and long prediction horizons.

Comparing the TCN and logistic regression baseline models in internal validation on the MIMIC-IV test set, TCN models consistently outperformed logistic regression models across all evaluation strategies and prediction horizons. In contrast, during external validation on BerlinICU without onset matching, logistic regression models approached or even exceeded the performance of TCN models at certain prediction horizons (Figure S2).

## Discussion

In this study, we assessed the impact of commonly used evaluation strategies – fixed horizon, peak score and continuous evaluation – on the performance of machine learning models for sepsis onset prediction. Our results demonstrate that the choice of the evaluation strategy significantly influences performance estimates, even though the same model is used on the same dataset.

We further addressed a gap in research involving the German healthcare system by aggregating an ICU dataset – called BerlinICU – from one of the largest university hospitals in Europe, Charité – Universitätsmedizin Berlin. Including more than 40,000 admissions from eight ICU wards, this dataset is one of the largest ICU datasets in Europe, exceeding the size of the currently available European datasets AUMCdb (24), HiRID-I (25) and SICdb (26). To the best of our knowledge, this is one of the first studies to explore how sepsis prediction models perform on large-scale German ICU data.

### Fixed horizon evaluation requires prospective knowledge of sepsis onset

Fixed horizon evaluation uses data up to a pre-defined time point before sepsis onset – the prediction horizon – to make predictions. Performance decreased with longer prediction horizons due to increasing temporal distance from sepsis onset, but remained at an overall high level. Without onset matching, performance estimates strongly increased for all horizons on the MIMIC-IV test set and for most horizons for BerlinICU, with the exception of the 6-hour horizon. This performance increase when not using onset matching is most parsimoniously explained by the evaluation of control patients over their entire stay, with emphasis on the period just before discharge. During these pre-discharge hours, control patients are typically in a stable condition, making it easier for the model to correctly identify them as low-risk for sepsis. This effect, also noted by Futoma (19), shows the significant impact of onset alignment. Despite this significant influence, many studies do not comment on the implementation of case-control matching (13). While the fixed horizon approach achieved the highest AUROC in a study on different evaluation strategies (18), we observed more nuanced performance differences compared to the other evaluation approaches depending on the dataset, horizon and onset matching approach. Generally, interpretation of performance metrics computed through the fixed horizon approach is challenging, as it relies on prospective information – the time point of the sepsis onset – which is not accessible in a real-world clinical setting.

### Peak score evaluation is influenced by the length of stay distribution and requires onset matching to avoid bias

The peak score evaluation assesses the model’s peak confidence in predicting sepsis throughout the entire ICU stay. This approach seems intuitively aligned with clinical practice, where a fixed threshold would be pre-determined, and an alarm is triggered once the prediction model’s output surpasses it — potentially just once per patient, with subsequent alarms suppressed (16, 20). The focus is on whether an alarm is raised at any point during the patient’s stay, which is replicated by determining the maximum prediction score. However, the performance metrics determined under this approach are influenced by differences in length of stay distributions between cases and controls: longer time series for control patients increase the likelihood of higher maximum prediction scores for these patients simply due to random fluctuations, ultimately reducing the AUROC as the maximum scores for control patients approach those of sepsis patients on average (Figure S4). Importantly, this bias conflicts with the core interpretation of the AUROC, which measures the probability that a randomly selected true positive will have a higher predicted score than a randomly selected true negative. Our results demonstrate this behavior, showing that performance estimates without onset matching – where control cases tend to have longer length of stays – were significantly reduced compared to analyses with onset matching (Figure 4). To mitigate this effect, an onset matching approach that aligns the length of stay distributions between positive and negative cases is strictly necessary when applying this evaluation strategy.

The discrepancy between the peak score evaluation across all patients (Figure 4), where onset matching increases the AUROC, and horizon-dependent analysis (Figure 5), where onset matching decreases the AUROC, can be explained by the exclusion of patients with insufficient stay lengths in the latter. This exclusion alters the length of stay distribution and thereby significantly impacts performance metrics. Not filtering patients with insufficient length of stays shows a distinct interaction effect between onset matching and prediction horizon (Figure S3), further highlighting the critical influence of length of stay distribution on the performance estimates in the peak score evaluation approach.

### Continuous evaluation without onset matching reflects clinical practice

Continuous prediction evaluation showed increased performance with shorter prediction windows, demonstrating the model’s effectiveness in utilizing short-term, relevant data to predict sepsis onset. As the prediction window narrows and focuses more on the actual onset of sepsis, the assigned sepsis labels align more closely with the sepsis-related signals in the patient data, enhancing prediction efficiency. Interestingly, although the models were trained to predict sepsis 6 hours in advance, this specific labeling window did not yield the best performance.

Evaluating on BerlinICU with onset matching, model performance was significantly reduced compared to the MIMIC-IV test set (ΔAUROC −0.11 for 6 hours before onset; −0.08 for 1 hour). This reduced performance when transferring models between datasets was also evident in a previous study where models trained on MIMIC-IV were evaluated on other European datasets using a horizon-based evaluation with a six hour window (17). Even though the performance in that study declined for European datasets (HiRID-I: −0.07, AUM-Cdb: −0.09) as well as for another US dataset (eICU: −0.10), it was not as severe as observed in BerlinICU.

Without onset matching, performance in the MIMIC-IV test set was higher across horizons, and was similar or better on BerlinICU for most horizons. Again, this higher performance without onset matching most likely reflects the inclusion of time points closer to discharge for control patients, who generally have a healthier profile, making the prediction task easier. While onset matching may provide a focused assessment in more challenging cases, not using it better mirrors clinical practice, where patients are monitored continuously from admission to discharge. In our view, continuous evaluation without onset matching better reflects real-world clinical application, as the AUROC represents the likelihood of a positive case being ranked higher than a negative one, including time points closer to discharge.

Although this approach requires pre-determining a time window, which indicates how far in the future the onset may be predicted, we consider continuous evaluation the better choice for two reasons: First, defining a time window — ideally based on pathophysiological plausibility — ensures that only alarms occurring within a clinically meaningful period before sepsis onset are considered. This distinction is crucial, as some published models show high performance days to weeks before the actual event (27–29), likely detecting broader, correlated health-related features rather than sepsis specifically. This underlines the importance of thoroughly defining and testing what these models are predicting to ensure their practical utility in clinical settings. Second, varying the prediction window and reporting the performance across all these windows allows for a more nuanced understanding of the models’ ability to predict event onset in advance.

### Limitations

Our study has several limitations: First, we did not have access to microbiological data, requiring an adjustment of the Sepsis-3 definition. This approach, however, was also chosen in other studies when using datasets from hospitals that did not provide microbiological data (16, 17). Second, we used models trained for a 6-hour horizon and applied them across various prediction horizons to predict sepsis onset. While these models may not perform optimally outside their original specifications, the inability to know the exact duration until a potential sepsis onset at any given timepoint in clinical practice necessitates using a flexible approach. Thus, by testing these models beyond their intended parameters, we can evaluate their robustness and generalizability to different clinical scenarios and timelines.

### Comparison with prior work

Our work combines the idea of generalizability across countries (16, 17) and suitable metric calculation concepts (16, 30). Our study builds on these previous concepts, refining and extending them as needed. While Moor and Rockenschaub examined the influence of various algorithms and training sets from different countries, our work focuses on different evaluation strategies. We applied sepsis onset matching (19, 30) and peak score evaluation (16), combining approaches that were presented separately. As the peak score evaluation is only suitable for retrospective analysis, we also applied a continuous evaluation strategy (13). Moreover, there are only few studies that have applied models to German data (29, 31), none of which having a dedicated focus on comparing performances in different evaluation strategies.

Several studies have applied sepsis prediction models prospectively to US data (14, 15, 32), but they often lack details on how the models were trained and how data was preprocessed. This lack of transparency is a common issue, as noted in two systematic reviews of sepsis prediction models address (12, 13). Accordingly, common checklists for reporting prediction models include items for external validation in different countries and time periods (33, 34).

### Conclusions

Our study highlights the critical need to choose an evaluation approach that is aligned with the intended interpretation of the resulting performance metric, as different approaches yield markedly different performance estimates despite using the same model on the same dataset. Importantly, for the fixed horizon and peak score evaluation approaches, a carefully devised onset matching strategy is crucial to avoid skewed results that may not reflect the true model performance. In our view, the continuous evaluation approach better reflects clinical reality. Despite requiring specifying the horizon time window, it offers insights into the model’s effectiveness in a clinical setting.

## Data Availability

Aggregated data produced in the present study are available upon reasonable request to the authors. Individual patient-level data can not be shared due to data privacy regulations.

## ACKNOWLEDGEMENTS

We thank Fabian Schreiber and Falk Meyer-Eschenbach for their support in data extraction. The authors also acknowledge the Scientific Computing of the IT Division at the Charité - Universitätsmedizin Berlin for providing computational resources that have contributed to the research results reported in this paper.

## Supplementary Appendix

### A. Features

This section lists the clinical features used in the prediction models, categorized into static variables, vital signs, arterial blood gas analysis, electrolytes, blood count, and other laboratory parameters.

**Supplementary Table S1.**
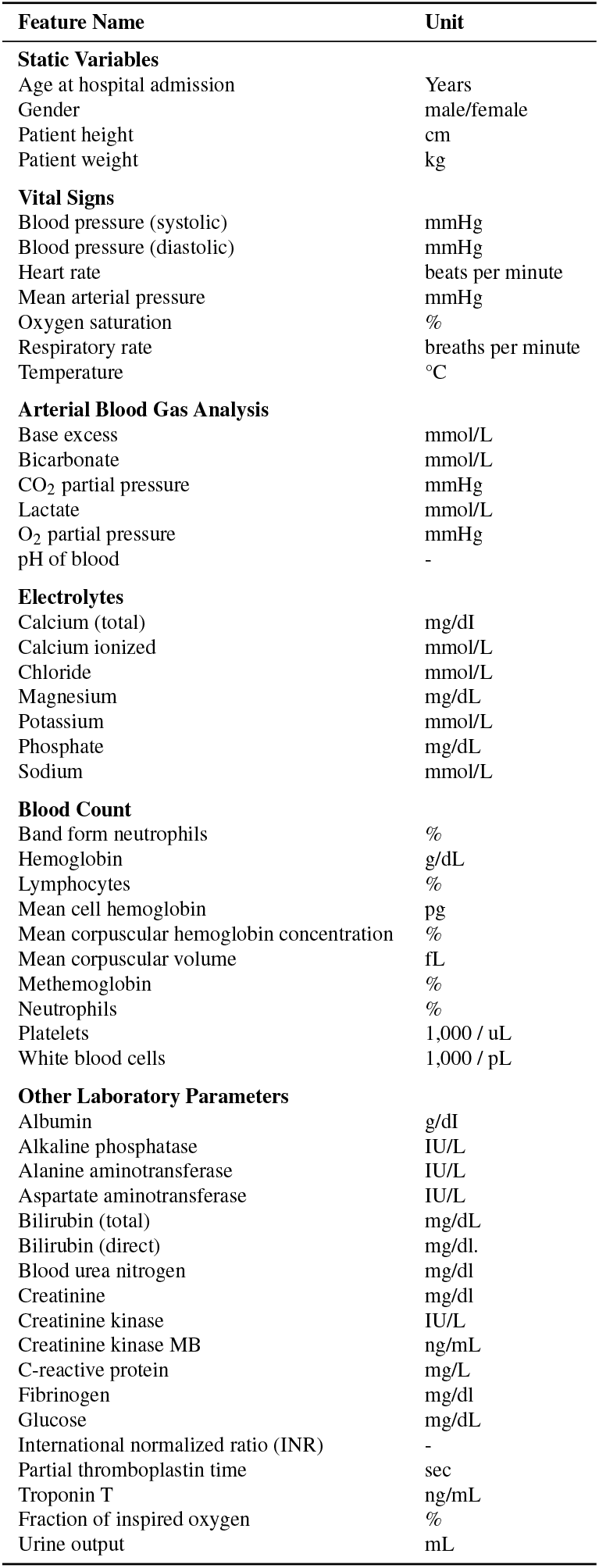
Clinical features used in the prediction models.

### B. Cohort Sizes

This section provides an overview of the dataset sizes across different prediction horizons, including the number of total patients, sepsis cases, and prevalence rates for MIMIC-IV and BerlinICU cohorts.

**Supplementary Table S2.** Cohort sizes for the time-dependent evaluation. Shown is the number of patients in each cohort and dataset after filtering for patients whose length of stay matches or exceeds the prediction horizon. For BerlinICU, the total cohort size refers to the complete dataset, while for MIMIC-IV the mean, min and max across all test sets is described (TCN: 10 test sets, LogReg: 5 test sets). TCN: Temporal Convolutional Network. LogReg: Logistic Regression.

| Dataset \| Horizon | Mean Total Patients (Min-Max) | Mean Sepsis Patients (Min-Max) | Mean Prevalence (%) (Min-Max) |
| --- | --- | --- | --- |
| **MIMIC-IV (TCN)** |  |  |  |
| Total | 13,411 (13,411- 13,411) | 747 (709-830) | 5.6 (5.3-6.2) |
| -100 | 1,664 (1,616-1,724) | 60 (45-69) | 3.6 (2.8-4.2) |
| -75 | 2,716 (2,613-2,783) | 101 (89-115) | 3.7 (3.3-4.3) |
| -48 | 5,184 (5,036-5,246) | 149 (132-179) | 2.9 (2.6-3.5) |
| -24 | 9,934 (9,882-9,990) | 225 (207-271) | 2.3 (2.1-2.7) |
| -12 | 12,686 (12,644-12,719) | 435 (395-500) | 3.4 (3.1-4.0) |
| -6 | 13,411 (13,411- 13,411) | 747 (709-830) | 5.6 (5.3-6.2) |
| -3 | 13,411 (13,411- 13,411) | 747 (709-830) | 5.6 (5.3-6.2) |
| -2 | 13,411 (13,411- 13,411) | 747 (709-830) | 5.6 (5.3-6.2) |
| -1 | 13,411 (13,411- 13,411) | 747 (709-830) | 5.6 (5.3-6.2) |
| **MIMIC-IV (LogReg)** |  |  |  |
| Total | 13,411 (13,411-13,412) | 746 (746-746) | 5.6 (5.6-5.6) |
| -100 | 1,676 (1,647-1,716) | 58 (45-67) | 3.4 (2.7-3.9) |
| -75 | 2,727 (2,685-2,800) | 96 (89-106) | 3.5 (3.3-3.8) |
| -48 | 5,203 (5,136-5246) | 146 (137-153) | 2.8 (2.6 – 2.9) |
| -24 | 9,926 (9,857-9,984) | 218 (204-231) | 2.2 (2.0 - 2.3) |
| -12 | 12,683 (12,664-12,703) | 431 (408-442) | 3.4 (3.2-3.5) |
| -6 | 13,411 (13,411-13,412) | 746 (746-746) | 5.6 (5.6-5.6) |
| -3 | 13,411 (13,411-13,412) | 746 (746-746) | 5.6 (5.6-5.6) |
| -2 | 13,411 (13,411-13,412) | 746 (746-746) | 5.6 (5.6-5.6) |
| -1 | 13,411 (13,411-13,412) | 746 (746-746) | 5.6 (5.6-5.6) |
| **BerlinICU** |  |  |  |
| Total | 40,155 | 4,137 | 10.3 |
| -100 | 4,357 | 504 | 11.6 |
| -75 | 6,277 | 773 | 12.3 |
| -48 | 9,897 | 1,242 | 12.6 |
| -24 | 18079 | 2,121 | 11.7 |
| -12 | 33,798 | 2,871 | 8.5 |
| -6 | 40,132 | 4134 | 10.3 |
| -3 | 40,132 | 4134 | 10.3 |
| -1 | 40,132 | 4134 | 10.3 |

### C. Baseline Models

This section presents the performance of logistic regression models across different evaluation strategies, including fixed horizon, peak score, and continuous evaluation, comparing results between MIMIC-IV and BerlinICU datasets.

**Supplementary Figure S1.**
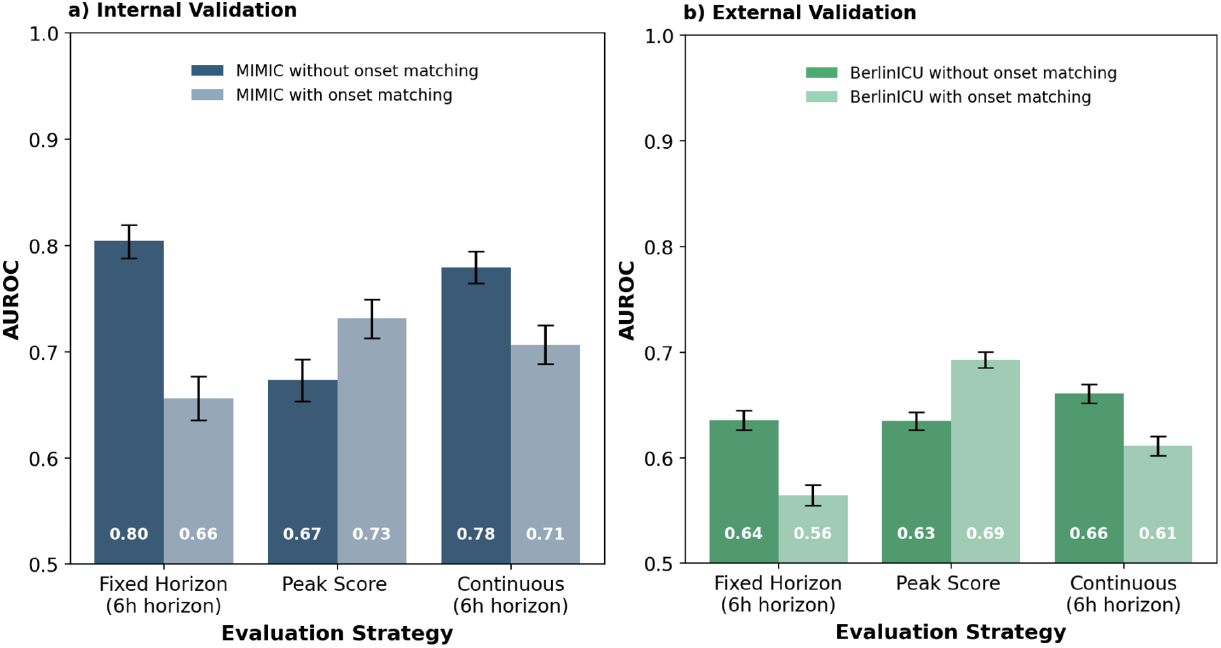
Comparison of model performance of logistic regression models for different evaluation strategies. The models were applied to the test splits of the MIMIC-IV training dataset (a) and the entire German ICU dataset (b; BerlinICU). Model performance was evaluated using fixed horizon evaluation (6h horizon), peak score evaluation (entire patient stay) and continuous evaluation (6h horizon) without onset matching (dark bars) and with onset matching (light bars). The AUROC value inside the bar represents the mean AUROC across all bootstrap samples and models. AUROC: area under the receiver operating characteristic. Error bars: 95% confidence intervals.

**Supplementary Figure S2.**
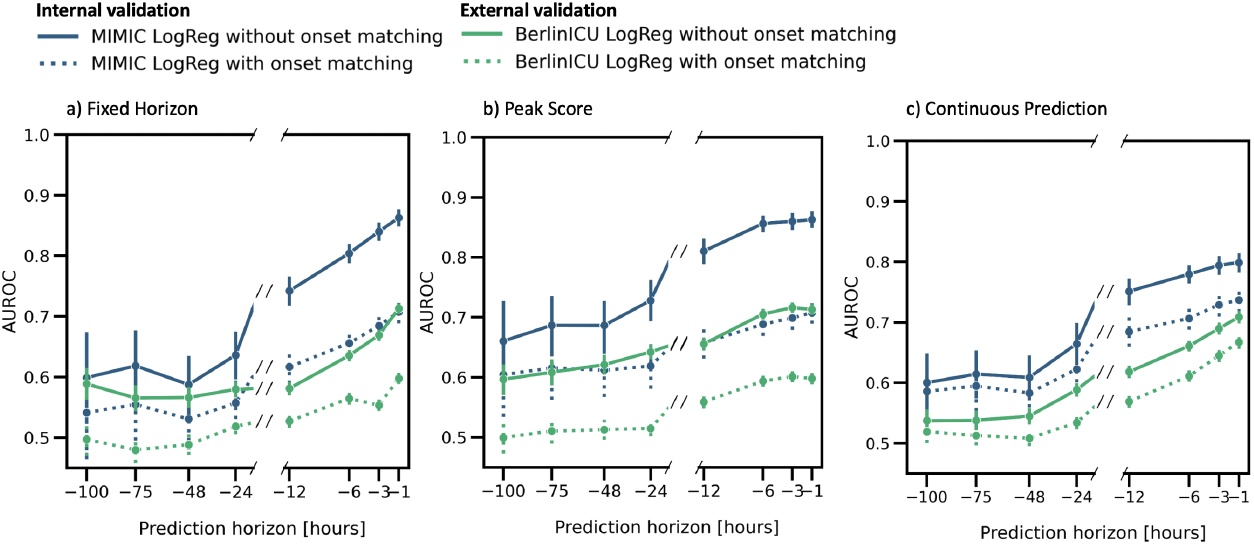
Performance metrics across prediction horizons for logistic regression models, comparing the performance an evaluation with onset matching (dashed lines) and without onset matching (solid lines). The models were applied to the test splits of the MIMIC-IV training dataset (blue) and the entire German ICU dataset (BerlinICU; green). Shown is the AUROC based on varying prediction horizons from one hour before onset up to 100 hours before onset. AUROC: area under the receiver operating characteristic. Error bars: 95% confidence intervals.

**Supplementary Figure S3.**
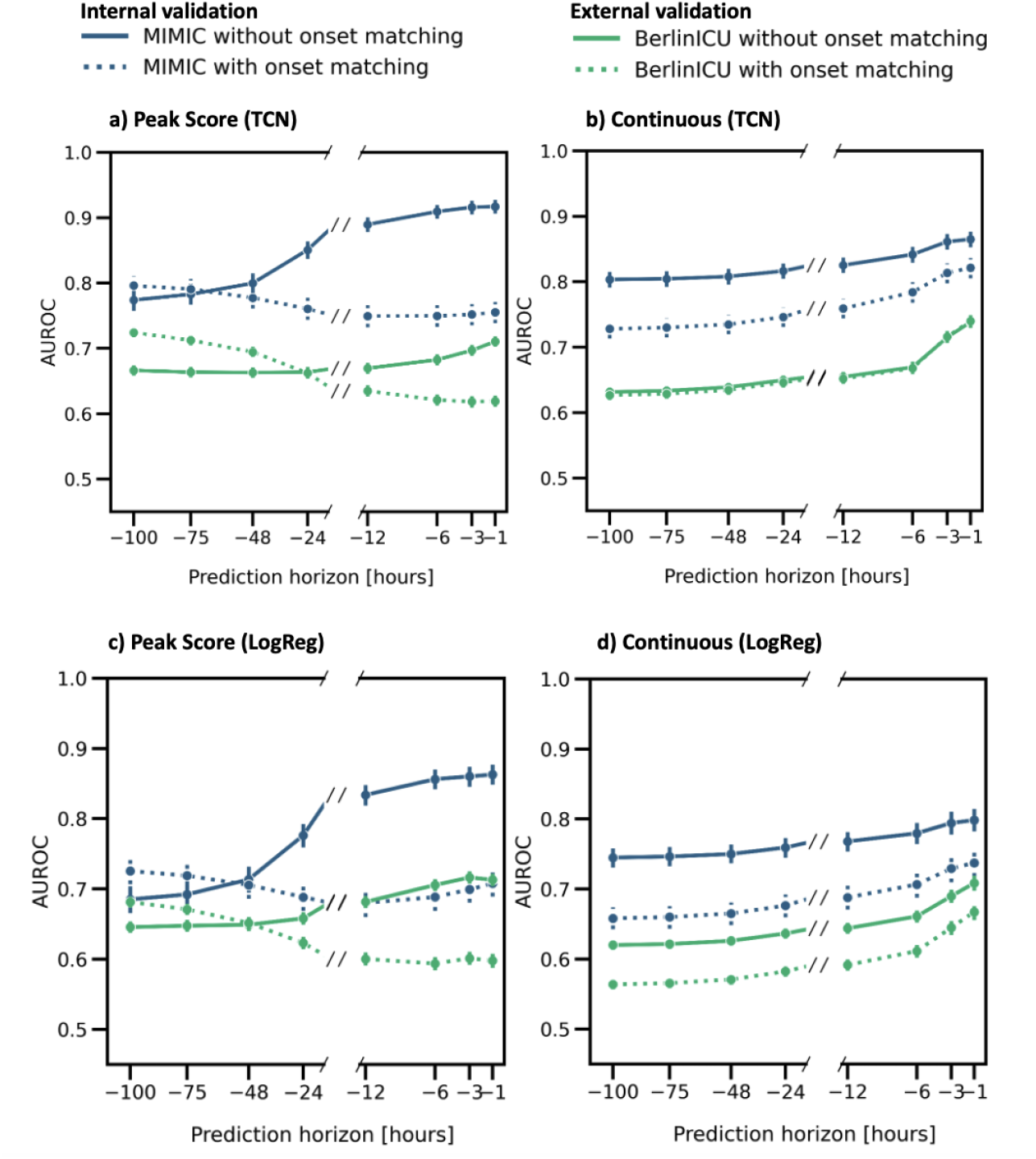
Performance metrics across prediction horizons, without filtering for length of stay (LOS), comparing, the performance of temporal convolutional network (TCN, a-b) and logistic regression (LogReg, c-d) models for an evaluation with onset matching (dashed lines) and without onset matching (solid lines). The models were applied to the test splits of the MIMIC-IV training dataset (blue) and the entire German ICU dataset (BerlinICU; green). Shown is the AUROC based on varying prediction horizons from one hour before onset up to 24 hours before onset. Each prediction horizon consisted of the same patients. If the LOS of a patient was shorter than the prediction horizon, then evaluation strategies were applied to the data points that were available. AUROC: area under the receiver operating characteristic. Error bars: 95% confidence intervals.

### D. Peak Score Evaluation Bias

This section illustrates the potential bias introduced by peak score evaluation using simulated random data, demonstrating how the number of samples per patient affects classifier performance, even when the underlying data distributions are identical.

**Supplementary Figure S4.**
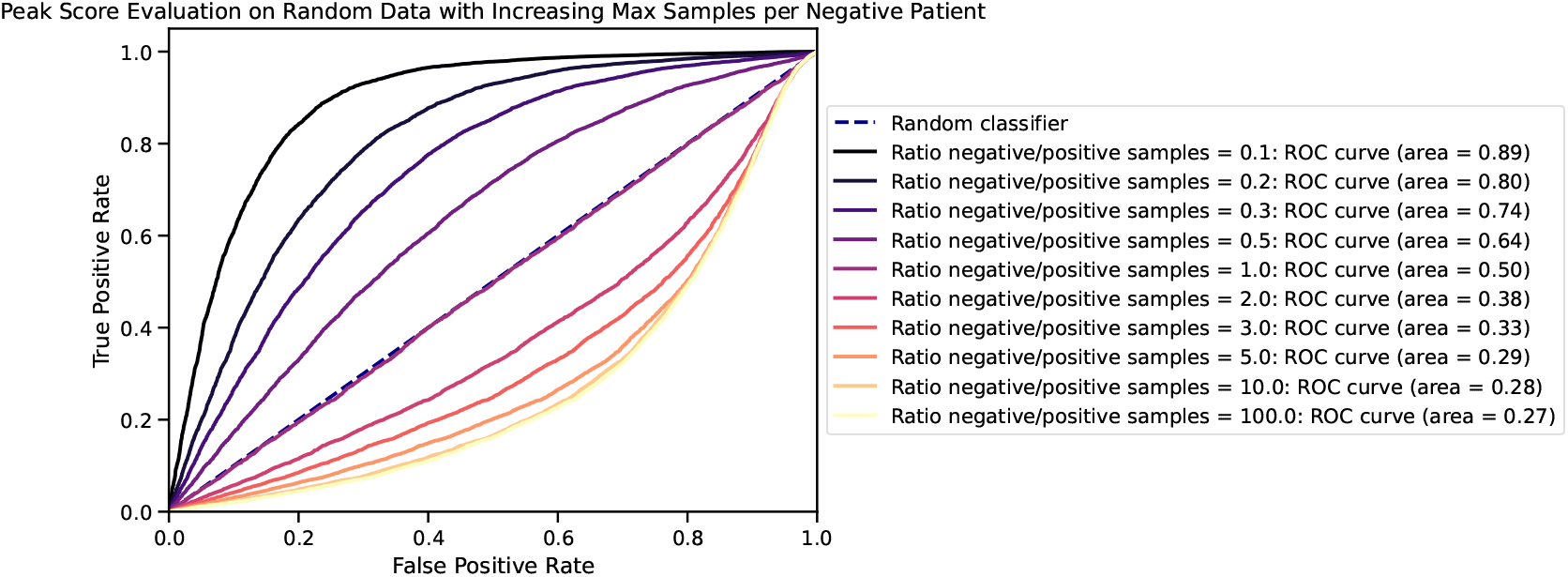
Peak Score Evaluation on Random Data with Increasing Maximum Number of Samples per Negative Patient. The figure illustrates the behavior of a classifier on simulated data where there is no inherent difference between the negative and positive patients. Both groups consist of 10,000 patients each, with scores drawn from the same random Gaussian distribution. The only distinction between the groups is that each positive patient has up to 10 samples, while the negative patients have varying maximum numbers of samples, as indicated in the legend. The results demonstrate that when taking the maximum score for each patient, the classifier metric becomes increasingly skewed as the number of samples for each negative patient deviates more from the number of samples per positive patient. This disparity between the number of samples per class creates an artificial boost or dilution in performance, even though there is no true difference between the underlying data distributions for the two groups. The diagonal line (“Random classifier”) shows the baseline performance of a random classifier, and the comparison highlights the inflation of the metric due to unequal sampling.

